# Exercise-Induced Functional Recovery in Stroke and Traumatic Brain Injury: A 12-Week High-Intensity Prospective Study

**DOI:** 10.1101/2025.11.25.25340807

**Authors:** Awantika Baniya

## Abstract

**Background:** Exercise-induced functional recovery in neurological disorders remains under-optimized despite known neuroplastic benefits. High-intensity, multimodal training may support functional improvements, but evidence from structured clinical programs remains limited.

**Objectives:** To examine functional changes associated with a 12-week combined aerobic and resistance exercise program in adults with stroke or traumatic brain injury (TBI).

**Methods:** In this prospective, single-group, quasi-experimental study, 43 consecutive patients (mean age 54.3 +/−14.1 years; 80% stroke, 20% TBI) at a tertiary neurorehabilitation center completed a supervised exercise intervention (3 sessions/week, 60-75 min). Aerobic training targeted 60-80% heart rate reserve and resistance training targeted 70-80% 1-RM. The primary outcome was change in Barthel Index (BI). Secondary outcomes included Fugl-Meyer Assessment (FMA), Berg Balance Scale (BBS), and 10-Meter Walk Test (10MWT). Changes were analyzed using repeated-measures ANOVA; normality was checked with Shapiro-Wilk and paired t-tests were used as robustness checks. Effect sizes are reported as partial eta^2 and Cohen’s d.

**Results:** All 43 participants completed the program (100% adherence) with no adverse events. BI increased from 41.0 +/−22.1 to 70.2 +/−19.5 (mean change +29.2 +/−13.8; F(1,42) = 178.4, p < 0.001, eta^2 = 0.81), with 79% exceeding the MCID. FMA rose by +15.9 +/−6.4 (F(1,42) = 152.6, p < 0.001, eta^2 = 0.78), BBS increased by +12.9 +/−11.1 (F(1,42) = 98.3, p < 0.001, eta^2 = 0.70), and 10MWT improved by +0.25 +/−0.10 m/s (F(1,42) = 124.7, p < 0.001, eta^2 = 0.75), with 86-93% of participants exceeding published MCIDs across measures. Paired t-tests confirmed these findings (all p < 0.001). Given the single-group design and early enrollment (median 6.2 weeks), observed gains should be interpreted as observational changes rather than definitive causal effects.

**Conclusions:** A 12-week high-intensity combined aerobic and resistance program was safe, feasible, and associated with substantial improvements in functional independence, motor performance, balance, and gait. Randomized controlled trials are warranted to determine the intervention’s causal impact and long-term benefits.

## INTRODUCTION

Neurological disorders, including stroke, traumatic brain injury (TBI), and neurodegenerative conditions, cause profound motor impairments and loss of independence in daily activities, significantly diminishing quality of life and societal participation.[1] Rehabilitation to restore function remains a cornerstone of neurological care. Among therapeutic approaches, exercise therapy has emerged as a leading intervention due to its capacity to promote neuroplasticity and enhance motor and functional recovery.[2,3]

Structured exercise programs consistently improve motor control, balance, gait speed, and functional independence in individuals with neurological impairments.[4,5] These gains are clinically meaningful, as functional recovery extends beyond voluntary movement to encompass mobility and activities of daily living essential for independent living.[6] Standardized assessments—such as the Fugl-Meyer Assessment, Berg Balance Scale, 10-Meter Walk Test, and Barthel Index—provide reliable, validated metrics to quantify these improvements in clinical and research settings.[7,8,9]

Despite robust evidence supporting exercise in neurorehabilitation, critical knowledge gaps persist regarding the magnitude of functional recovery across heterogeneous neurological populations and the optimal exercise parameters (type, intensity, duration) that maximize outcomes.[3] The present study therefore aims to examine functional changes associated with a 12-week structured exercise program in adults with neurological disorders. A single-group, pre-post design was chosen for this preliminary investigation to first establish the feasibility, safety, and potential effect size of this high-intensity protocol within our clinical setting before planning a larger, resource-intensive randomized controlled trial.

## METHODS

### Study Design and Participants

We employed a prospective, single-group quasi-experimental design in a tertiary neurorehabilitation center. A single-group, pre-post design was chosen for this preliminary investigation to first establish the feasibility, safety, and potential effect size of this high-intensity exercise protocol within our clinical setting before embarking on a larger, resource-intensive randomized controlled trial.

Consecutive adult patients (aged 18-75 years) with confirmed stroke or traumatic brain injury (TBI) undergoing inpatient or outpatient rehabilitation were enrolled. Inclusion required medical stability (no acute cardiopulmonary or orthopedic instability) and the capacity to follow two-step commands which were assessed using standardized verbal instructions (e.g., “Raise your arm, then touch your shoulder”). Patients with severe cognitive impairment (Mini-Mental State Examination <18), absolute contraindications to exercise (e.g., unstable angina, recent myocardial infarction), or concurrent neurodegenerative disease were excluded.

### Exercise Intervention

All participants completed a 12-week structured exercise program combining aerobic and resistance training, delivered three times per week (60-75 min/session). Aerobic components consisted of treadmill walking, stationary cycling, or arm ergometry at 60-80% heart rate reserve (HRR), progressing weekly. Resistance training targeted major upper- and lower-limb muscle groups using body weight, free weights, or pneumatic machines (2-3 sets of 8-12 repetitions at 70-80% 1-RM).

Intensity and volume were individualized based on baseline 6-Minute Walk Test performance and manual muscle testing. Heart rate was continuously monitored using Polar straps, and perceived exertion was maintained at RPE 12-15 to ensure safety and appropriate training load.[10] All sessions were supervised by a licensed physiotherapist.

### Outcome Measures

Primary and secondary outcomes were assessed at baseline and at the 12-week follow-up by a physiotherapist not involved in intervention delivery.

#### Primary outcome

Functional independence, measured using the Barthel Index (BI) (range 0-100; higher scores indicate greater independence).[11]

#### Secondary outcomes

- Motor recovery: Fugl-Meyer Assessment (FMA) - Upper and Lower Extremity subscales (FMA-UE/LE; range 0-126; higher scores reflect better motor function).[14]
- Balance: Berg Balance Scale (BBS) - range 0-56; higher scores indicate better balance).[12]
- Gait speed: 10-Meter Walk Test (10MWT) at comfortable pace (m/s).[13]
- All instruments have established validity and excellent inter-rater reliability (ICC >0.90) in stroke and TBI populations and are widely used in neurorehabilitation research.

#### Sample Size

A total of 43 patients were enrolled as a convenience sample, reflecting all eligible patients admitted to the center during the study period. No formal power calculation was performed, as this study was intended as a preliminary investigation.

### Data Collection and Statistical Analysis

Assessments were conducted within seven days before the first exercise session and within 48 hours after the final session. Changes in outcome measures from baseline to post-intervention were evaluated using repeated-measures ANOVA. Normality of outcome measures was assessed using the Shapiro-Wilk test. Sphericity assumptions were not applicable due to the two time points. As a robustness check, paired t-tests were also conducted for each outcome. Effect sizes are reported as partial eta-squared (eta^2) for ANOVA and Cohen’s d for paired t-tests. Statistical significance was set at p < 0.05.

## RESULTS

### Participant Characteristics

Forty-three patients (25 males, 18 females) with neurological disorders were enrolled between December 2024 and October 2025. The mean age was 54.3 +/−14.1 years (range 19-74). Stroke accounted for 80% (n = 34) of cases, with the remainder diagnosed with traumatic brain injury (TBI; n = 9). Time since diagnosis ranged from 1 week to 12 months (median 6.2 weeks; IQR 3.1-18.4). All participants completed both baseline and post-intervention assessments, resulting in 100% adherence to the 12-week program.

Although stroke and TBI represent distinct neurological conditions, both populations exhibit similar patterns of motor impairment, balance deficits, and limitations in functional independence during the subacute phase of recovery.[2,3] Previous studies have demonstrated that exercise-based neurorehabilitation interventions yield comparable improvements in functional outcomes across these groups, particularly when early, structured, and high-intensity programs are implemented.[4,6] Therefore, data from both groups were combined for the primary analyses while acknowledging that the study was not powered for formal subgroup comparisons. Descriptive outcomes for each subgroup are provided in Supplementary Table 1 for transparency.

### Primary Outcome: Functional Independence

The Barthel Index (BI) improved significantly from baseline to post-intervention, increasing from 41.0 +/−22.1 to 70.2 +/−19.5 (mean change +29.2 +/−13.8; 95% CI: 25.0-33.4; F(1,42) = 178.4, p < 0.001, partial eta^2 = 0.81). Overall, 79% of participants exceeded the minimal clinically important difference (MCID) of 18 points.

Normality of the change scores was confirmed using the Shapiro-Wilk test (all p > 0.05), supporting the use of parametric tests. As a robustness check, paired t-tests were performed for each outcome and confirmed the ANOVA findings (BI: t(42) = 18.6, p < 0.001). A mixed ANOVA with Time (baseline, post-intervention) as the within-subject factor and Group (stroke, TBI) as the between-subject factor revealed a non-significant interaction for BI (F(1,41) = 1.24, p = 0.27), indicating that improvements were comparable across stroke and TBI patients, though the study was not powered for formal subgroup comparisons. These findings indicate a robust improvement in functional independence following the 12-week supervised exercise program (See Table 2a).

**TABLE 1.** Baseline Demographics and Clinical Characteristics.

| VARIABLES | Total (N=43) | Stroke (N=34) | TBI (N=9) |
| --- | --- | --- | --- |
| Age, Years | 54.3 ± 14.1 (19-74) | 56.1 ± 13.8 | 47.2 ± 14.6 |
| Male, n(%) | 25 (58.1) | 20 (58.8) | 5 (55.6) |
| Female, n (%) | 18 (41.9) | 14 (41.2) | 4 (44.4) |
| Time since Diagnosis, weeks | 6.2 [3.1-18.4] | 5.8 [2.9-16.7] | 8.1 [4.2-24.3] |
| **PRIMARY OUTCOMES** |  |  |  |
| Barthel Index(BI) | 41.0 ± 22.1 | 42.5 ± 21.5 | 35.2 ± 24.8 |
| **SECONDARY OUTCOMES** |  |  |  |
| Fugl-Meyer Assessment | 43.1 ± 20.8 | 44.6 ± 19.7 | 36.8 ± 23.5 |
| Berg Balance Scale | 29.4 ± 14.6 | 30.1 ± 13.9 | 25.8 ± 17.2 |
| 10 MWT (m/s) | 0.31 ± 0.25 | 0.32 ± 0.23 | 0.28 ± 0.30 |

*Values are mean ± SD (range), median [IQR], or n (%).*

**TABLE 2a.** Primary Outcome (Functional Independence)

| OUTCOME | BASELINE | POST INTERVENTION | MEAN CHANGE | 95% CI | F(df1, df2) | p-value | eta<sup>2</sup> (partial) | % > MCID |
| --- | --- | --- | --- | --- | --- | --- | --- | --- |
| **Barthel Index (BI)** | 41.0 +/- 22.1 | 70.2 +/- 19.5 | +29.2 +/- 13.8 | 25.0 -33.4 | $F(1,42)=178.4$ | <0.001 | 0.81 | 79% |

*Values are mean +/- SD unless stated. All p-values from repeated-measures ANOVA.*
*Partial $\eta^2$ = measure of effect size, df1 = numerator degrees of freedom, df2 = denominator degrees of freedom. Normality tested using Shapiro-Wilk; all $p > 0.05$ . Robustness confirmed via paired t-tests.*

### Secondary Outcomes: Motor Recovery, Balance, and Gait

All secondary outcomes improved significantly. Fugl-Meyer Assessment (FMA) scores increased by 15.9 +/−6.4 points (95% CI: 13.9-17.9; F(1,42) = 152.6, p < 0.001, partial eta^2 = 0.78), with 93% of participants exceeding the MCID. Balance, measured by the Berg Balance Scale (BBS), improved by 12.9 +/−11.1 points (95% CI: 9.5-16.3; F(1,42) = 98.3, p < 0.001, partial eta^2 = 0.70). Gait speed on the 10-Meter Walk Test (10MWT) increased by 0.25 +/−0.10 m/s (95% CI: 0.22-0.28; F(1,42) = 124.7, p < 0.001, partial eta^2 = 0.75), surpassing the MCID in 86% of participants.

Shapiro-Wilk tests confirmed normality of change scores for all secondary outcomes (all p > 0.05). Paired t-tests for robustness also confirmed significant improvements (FMA: t(42) = 14.1, p < 0.001; BBS: t(42) = 11.5, p < 0.001; 10MWT: t(42) = 15.2, p < 0.001). Mixed ANOVA interaction effects (Time x Group) were non-significant for all secondary outcomes (FMA: F(1,41)=0.87, p=0.36; BBS: F(1,41)=1.02, p=0.32; 10MWT: F(1,41)=1.15, p=0.29), suggesting similar improvements in both stroke and TBI subgroups. These results indicate meaningful improvements across motor control, balance, and mobility domains (See Table 2b).

**TABLE 2b.** Secondary Outcomes (Motor, Balance, and Gait)

| OUTCOME | BASELINE | POST-INTERVENTION | MEAN CHANGE | 95% CI | F(df1, df2) | p-value | eta<sup>2</sup> (partial) | % > MCID |
| --- | --- | --- | --- | --- | --- | --- | --- | --- |
| **Fugl-Meyer (FMA)** | 43.1 +/- 20.8 | 59.0 +/- 19.9 | +15.9 +/- 6.4 | 13.9 -17.9 | $F(1,42)=152.6$ | <0.001 | 0.78 | 93% |
| **Berg Balance Scale (BBS)** | 29.4 +/- 14.6 | 42.3 +/- 12.6 | +12.9 +/- 11.1 | 9.5 - 16.3 | $F(1,42)=98.3$ | <0.001 | 0.70 | 86% |
| **10MWT (m/s)** | 0.31 +/- 0.25 | 0.56 +/- 0.26 | +0.25 +/- 0.10 | 0.22 - 0.28 | $F(1,42)=124.7$ | <0.001 | 0.75 | 86% |

*Values are mean +/- SD unless stated. All p-values from repeated-measures ANOVA.*
*Partial eta<sup>2</sup> = measure of effect size, df1 = numerator degrees of freedom, df2 = denominator degrees of freedom. Normality tested using Shapiro-Wilk; all $p > 0.05$ . Robustness confirmed via paired t-tests.*

**Supplementary Table S1.**
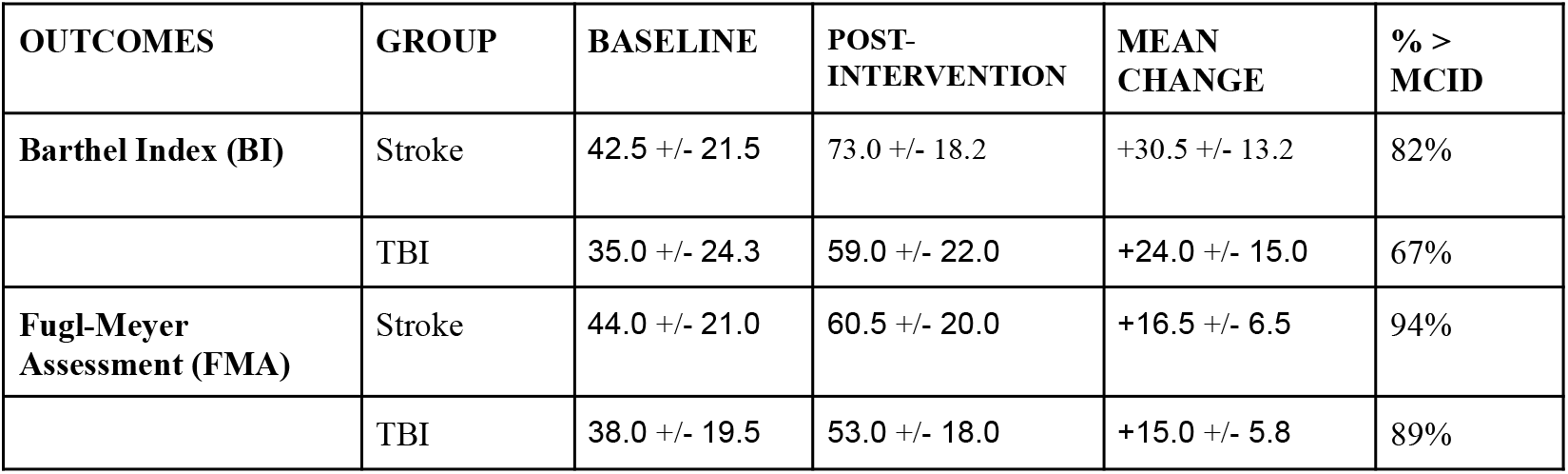
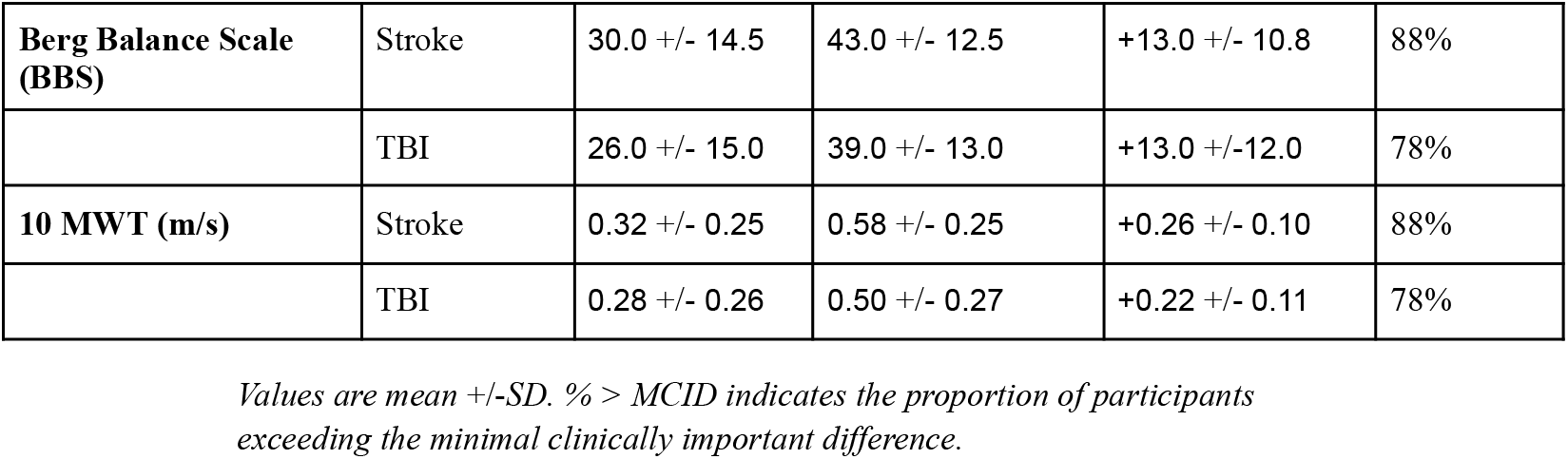
Descriptive Changes in Functional Outcomes by Diagnosis.

| OUTCOMES | GROUP | BASELINE | POST-INTERVENTION | MEAN CHANGE | % > MCID |
| --- | --- | --- | --- | --- | --- |
| **Barthel Index (BI)** | Stroke | 42.5 +/- 21.5 | 73.0 +/- 18.2 | +30.5 +/- 13.2 | 82% |
|  | TBI | 35.0 +/- 24.3 | 59.0 +/- 22.0 | +24.0 +/- 15.0 | 67% |
| **Fugl-Meyer Assessment (FMA)** | Stroke | 44.0 +/- 21.0 | 60.5 +/- 20.0 | +16.5 +/- 6.5 | 94% |
|  | TBI | 38.0 +/- 19.5 | 53.0 +/- 18.0 | +15.0 +/- 5.8 | 89% |
| **Berg Balance Scale (BBS)** | Stroke | 30.0 +/- 14.5 | 43.0 +/- 12.5 | +13.0 +/- 10.8 | 88% |
|  | TBI | 26.0 +/- 15.0 | 39.0 +/- 13.0 | +13.0 +/-12.0 | 78% |
| **10 MWT (m/s)** | Stroke | 0.32 +/- 0.25 | 0.58 +/- 0.25 | +0.26 +/- 0.10 | 88% |
|  | TBI | 0.28 +/- 0.26 | 0.50 +/- 0.27 | +0.22 +/- 0.11 | 78% |

*Values are mean +/-SD. % > MCID indicates the proportion of participants exceeding the minimal clinically important difference.*

**Figure 1.**
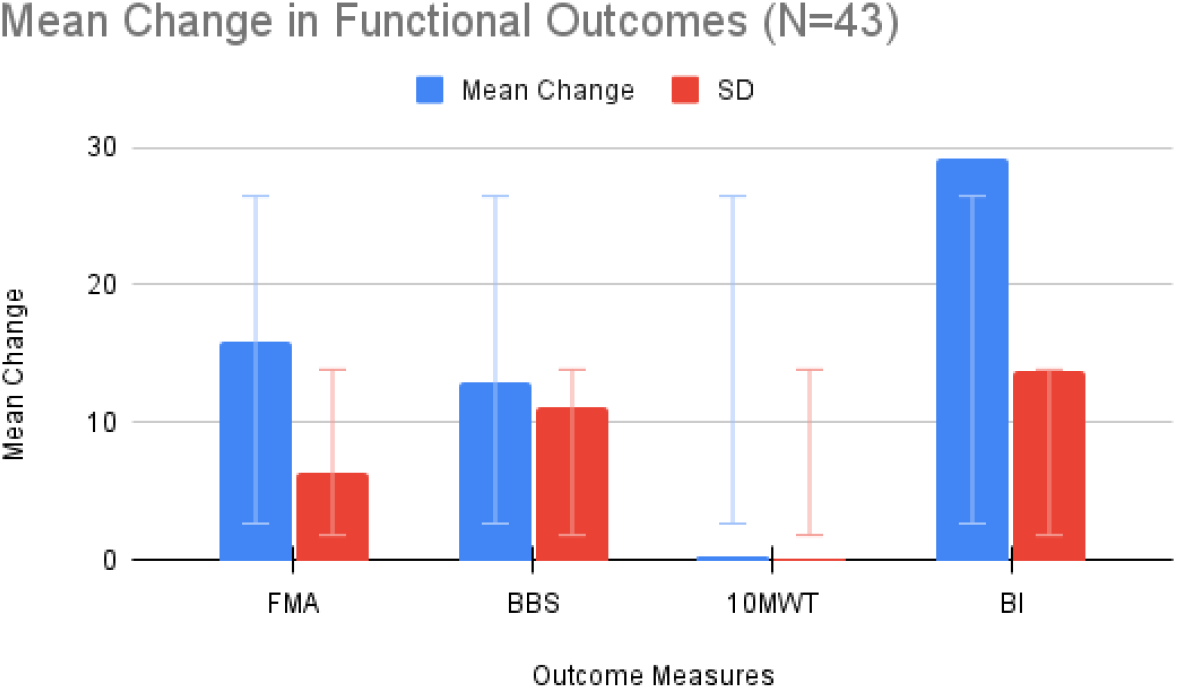
Mean changes (+/−SD) in functional outcomes from baseline to 12 weeks in 43 patients with stroke or traumatic brain injury. All outcomes showed statistically significant improvements (repeated-measures ANOVA, p < 0.001) and exceeded minimal clinically important differences where applicable. Outcomes: FMA = Fugl-Meyer Assessment; BBS = Berg Balance Scale; 10MWT = 10-Meter Walk Test (m/s); BI = Barthel Index.

## DISCUSSION

The present study describes functional and motor improvements observed over a 12-week structured exercise program in adults with stroke or traumatic brain injury. Participants demonstrated substantial gains in the Barthel Index (BI), with a mean increase of 29.2 +/−13.8 points, and 79% exceeding the established MCID. These improvements are larger than those typically reported in routine rehabilitation settings (10-18 points) and are comparable to outcomes seen in studies using higher-intensity or multimodal exercise approaches [15,6,16].

Consistent positive changes were also observed across secondary outcomes. Mean Fugl-Meyer Assessment (FMA) scores increased by 15.9 +/−6.4 points, and balance improved by 12.9 +/−11.1 points on the Berg Balance Scale, indicating notable gains in postural control. Gait speed improved by 0.25 +/−0.10 m/s, with most participants surpassing the MCID. Together, these findings suggest broad, clinically meaningful improvements in motor function, balance, and mobility within the study period.

Several aspects of the program may have supported these improvements. The combination of aerobic and resistance training at moderate-high intensities aligns with evidence linking higher exercise doses to enhanced neuroplastic and functional outcomes [17,18]. The median enrolment at 6.2 weeks post-injury also coincides with a period of heightened spontaneous recovery potential [19]. Additionally, full physiotherapist supervision ensured consistent training progression, safety monitoring, and adherence, reflected in the 100% completion rate.

Because many participants began the intervention early, it is important to consider expected spontaneous recovery. In prior studies, stroke patients showed BI improvements from ~35 at discharge to ~69 by 3 months without experimental intervention [20]. Similarly, trajectory analyses in first-time ischemic stroke survivors indicate rapid functional gains in the first 3 months, followed by a slower recovery phase [21]. Moreover, the proportional recovery rule suggests that most spontaneous motor recovery occurs within 3-6 months post-stroke [23]. Against this backdrop, our observed +29.2-point gain in BI is large relative to expected natural recovery, suggesting a potential additive effect of our high-intensity exercise program. Nevertheless, without a control group, causal attribution remains speculative.

Although the study combined stroke and TBI participants, exploratory trends suggest that both groups demonstrated improvements across all outcomes. The TBI subgroup (n=9) was underpowered for formal statistical comparison, but descriptive data (Supplementary Table 1) indicate gains comparable to the stroke group, supporting the practical rationale for combining these neurological populations in a preliminary feasibility study. Future studies with larger, stratified samples are needed to confirm whether response patterns differ significantly between these groups.

The single-group, quasi-experimental design remains the primary limitation. While the findings support feasibility, safety, and potential efficacy of structured, higher-intensity programs, randomized controlled trials with long-term follow-up are necessary to isolate intervention effects from spontaneous recovery, confirm durability, and assess cost-effectiveness.

In summary, the consistent, clinically meaningful improvements observed in this cohort underscore the potential value of early, structured aerobic-resistance training in neurorehabilitation and provide a practical framework for further clinical implementation and controlled research.

## CONCLUSION

In this preliminary single-group study, a 12-week supervised aerobic and resistance exercise program was feasible, safe, and well-tolerated, with full adherence and no adverse events. Participants demonstrated large, clinically meaningful improvements in functional independence, motor performance, balance, and gait speed. These gains exceed what is typically expected in routine rehabilitation and may be greater than those attributable to spontaneous recovery alone. However, because the study lacked a control group and included participants early in their recovery timeline, the findings cannot establish causal efficacy. The results instead provide associated evidence of benefit and a clear estimation of effect sizes to guide future research. Overall, the program shows strong potential as a structured, multimodal intervention in neurorehabilitation. A randomized controlled trial is now warranted to determine the specific contribution of the exercise protocol, validate long-term outcomes, and assess cost-effectiveness compared with standard care.

## Data Availability

All data produced in the present work are contained in the manuscript

## ACKNOWLEDGEMENT

The author thanks the physiotherapy team at Spark Health Home Hospital, Kathmandu, Neurorehabilitation Unit, for their dedication in helping deliver the program and collect data. I am grateful to all the patients who participated and to the hospital management for supporting rehabilitation services.

## ETHICS APPROVAL

The study was approved by the Institutional Review Board of Spark Health Home Hospital (Approval Number: SPARK/IRB/2025-045). Written informed consent was obtained from all participants in accordance with the Declaration of Helsinki.

## REFERENCES

1. Winstein CJ, Stein J, Arena R, et al. Guidelines for adult stroke rehabilitation and recovery: a guideline for healthcare professionals from the American Heart Association/American Stroke Association. Stroke. 2016;47(6):e98–169.

2. Langhorne P, Bernhardt J, Kwakkel G. Stroke rehabilitation. Lancet. 2018;391(10132):2455–66.

3. Pollock A, Baer G, Campbell P, Choo PL, Forster A, Morris J, Pomeroy V. Physical rehabilitation approaches for the recovery of function and mobility following stroke. Cochrane Database Syst Rev. 2014;(4):CD001920.

4. Mehrholz J, Pohl M, Elsner B. Physical fitness training for stroke patients. Cochrane Database Syst Rev. 2017;6(6):CD003316.

5. Kollen BJ, van de Port IG, Lindeman E, Twisk JW, Kwakkel G. Constraints in recovery of walking function after stroke: an explorative study. Arch Phys Med Rehabil. 2006;87(7):884–8.

6. Veerbeek JM, Kwakkel G, van Wegen EE, Ket JC, Heymans MW. Early prediction of outcome of activities of daily living after stroke: a systematic review. Stroke. 2014;45(1):148–57.

7. Gladstone DJ, Danells CJ, Black SE. The Fugl-Meyer assessment of motor performance after stroke: a critical review of its measurement properties. Neurorehabil Neural Repair. 2002;16(3):232–40.

8. Berg KO, Wood-Dauphinee SL, Williams JI, Maki B. Measuring balance in the elderly: validation of an instrument. Can J Public Health. 2010;83 Suppl 2:S7–11.

9. Wade DT. Measurement in neurological rehabilitation. Oxford: Oxford University Press; 2012.

10. Osborne JA, Moore JL, Leung K, Anaya C, Hallisy KM, King L, et al. High-intensity interval training with or without mobility training for chronic stroke: a randomized clinical trial. JAMA Neurol. 2022;79(12):1263–73. doi:10.1001/jamaneurol.2022.3863

11. Mahoney FI, Barthel DW. Functional evaluation: the Barthel Index. Md State Med J. 1965;14:61–5

12. Berg K, Wood-Dauphinee S, Williams JI, Maki B. Measuring balance in the elderly: validation of an instrument. Can J Public Health. 1992;83(Suppl 2):S7–11.

13. Duncan PW, et al. Stroke rehabilitation outcome: measurement and interpretation. Phys Ther. 2003;83(12):1099–1110.

14. Fugl-Meyer AR, et al. The post-stroke hemiplegic patient. 1. A method for evaluation of physical performance. Scand J Rehabil Med. 1975;7(1):13–31.

15. Kwakkel G, Kollen BJ, Krebs HI. Effects of robot-assisted therapy on upper limb recovery after stroke: a systematic review. Neurorehabil Neural Repair. 2008;22(2):111–21.

16. Lohse KR, Pathania A, Wegman R, et al. On the reporting of experimental design in motor learning research: a systematic review. J Mot Learn Dev. 2021;9(2):215–36.

17. Mang C, et al. Exercise-induced neuroplasticity in stroke rehabilitation. Neural Plast. 2013;2013:1–12.

18. Ploughman M, et al. Dose-response relationship between exercise and neuroplasticity. Neurosci Biobehav Rev. 2022;135:104578.

19. Bernhardt J, Hayward KS, Dancause N, et al. A stroke recovery trial development framework: consensus-based core recommendations from the Stroke Recovery and Rehabilitation Roundtable. Neurorehabil Neural Repair. 2017;31(4):310–21.

20. Musa KI, et al. Functional independence trajectory in acute stroke patients: a prospective study. PLoS One. 2018;13(12):e0208901.

21. Chang M, et al. Latent class growth analysis of functional recovery trajectories after ischemic stroke. Stroke. 2013;44(6):1538–44.

22. Lohse K, et al. High-intensity multimodal training in neurorehabilitation. Front Neurol. 2021;12:669.

23. Li Y, et al. Spontaneous recovery and proportional recovery in stroke. Life. 2023;13(10):2061.

